# Ensemble learning for higher diagnostic precision in schizophrenia using peripheral blood gene expression profile

**DOI:** 10.1101/2023.02.11.23285788

**Authors:** Vipul Vilas Wagh, Suchita Agrawal, Shruti Purohit, Tejaswini Pachpor, Leelavati Narlikar, Vasudeo Paralikar, Satyajeet Khare

## Abstract

The need for molecular biomarkers for schizophrenia has been well recognized. Peripheral blood gene expression profiling and machine learning (ML) tools have recently become popular for biomarker discovery. The stigmatization associated with schizophrenia advocates the need for diagnostic models with higher precision. In this study, we propose a strategy to develop higher-precision ML models using ensemble learning. We performed a meta-analysis using peripheral blood expression microarray data. The ML models, support vector machines (SVM), and prediction analysis for microarrays (PAM) were developed using differentially expressed genes as features. The ensemble of SVM-radial and PAM predicted test samples with a precision of 81.33% (SD: 0.078). The precision of the ensemble model was significantly higher than SVM-radial (63.83%, SD: 0.081) and PAM (66.89%, SD: 0.097). The feature genes identified were enriched for biological processes such as response to stress, response to stimulus, regulation of the immune system, and metabolism of organic nitrogen compounds. The network analysis of feature genes identified *PRF1, GZMB, IL2RB, ITGAL*, and *IL2RG* as hub genes. Additionally, the ensemble model developed using microarray data classified the RNA-Sequencing samples with moderately high precision (72.00%, SD: 0.08). The pipeline developed in this study allows the prediction of a single microarray and RNA-Sequencing sample. In summary, this study developed robust models for clinical application and suggested ensemble learning for higher diagnostic precision in psychiatric disorders.

**Research highlights:**

- Ensemble learning of Support Vector Machines (SVM) and Prediction Analysis for Microarrays (PAM) algorithms classified schizophrenia samples with higher precision.
- The pipeline developed in this analysis produced robust models with the ability to classify single microarray sample.
- Cross-platform validation of ensemble model using RNA-Sequencing data resulted in high precision.

**Graphical abstract:** 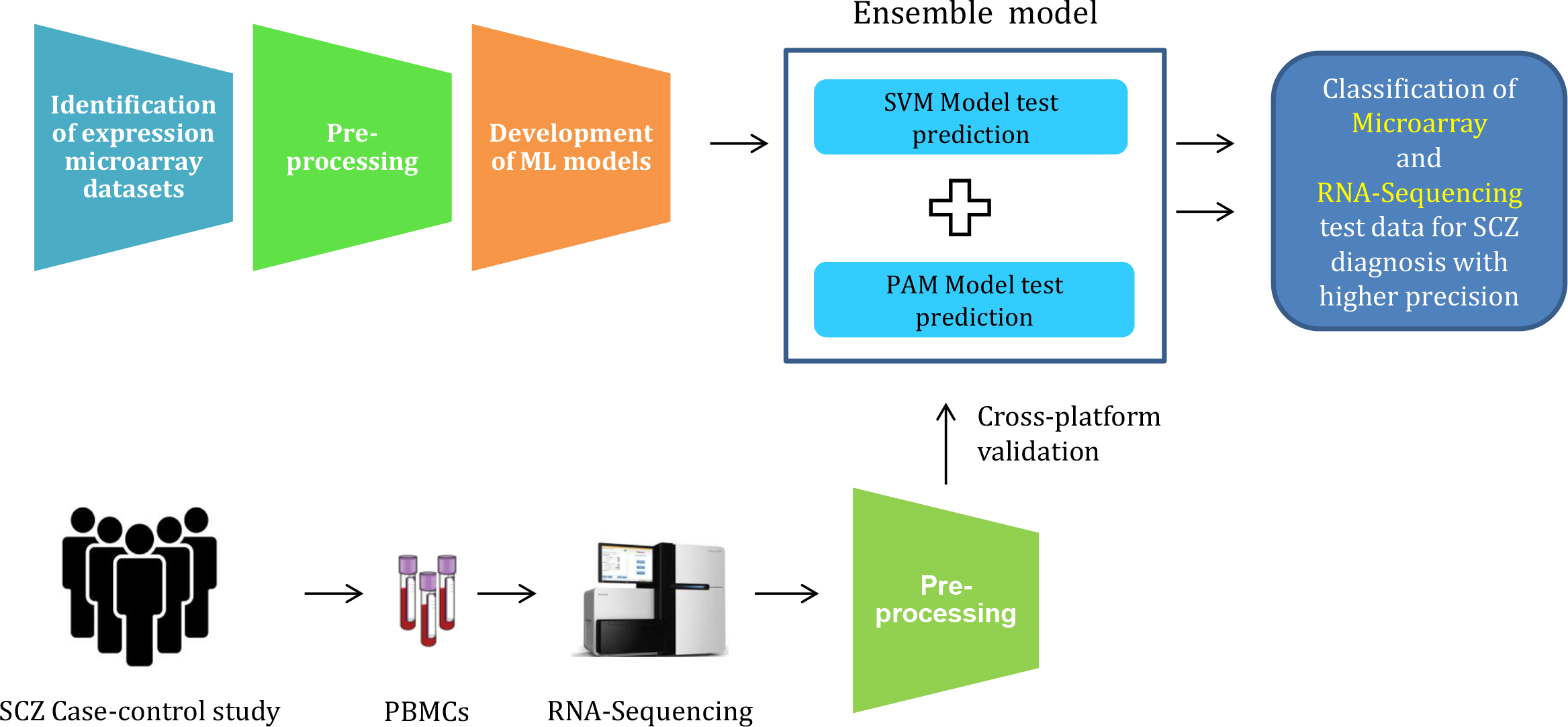

Blood based SCZ diagnosis using ensemble learning for higher precision

## 1. Introduction

Schizophrenia (SCZ) is a complex neuropsychiatric disorder characterized by a disruption in thinking and sense of self. The death rate is two times higher in schizophrenic patients, with cardiovascular diseases and suicide as the leading causes of death (Costa E Silva, 1998; Hennekens et al., 2005; Laursen et al., 2014). The lifetime prevalence of 0.2 - 0.4 % globally (GBD 2017 Disease and Injury Incidence and Prevalence Collaborators, 2018) indicates its universal presence irrespective of cultural differences worldwide. One of the significant issues in treating psychiatric disorders is delayed diagnosis. The current diagnostic procedure for SCZ is based on psychological evaluation, making it clinician dependent. The Diagnostic and Statistical Manual of Mental Disorders (DSM-5) based diagnosis for SCZ requires symptoms to be persistent for six months or more (American Psychiatric Association., 2013). The delay in the treatment accounts for a higher degree of years lived with disability associated with SCZ (GBD 2017 Disease and Injury Incidence and Prevalence Collaborators, 2018). Recent studies suggest the treatment outcome could be improved if the time elapsed before the treatment is reduced (Chan et al., 2014). Thus, having a blood test can surely strengthen and quicken the current diagnostic process for SCZ.

Molecular alterations such as gene expression changes associated with the disorder have been proposed to be used as potential biomarkers. Our previous study provides substantial evidence for using peripheral blood gene expression profiles for biomarker discovery (Wagh et al., 2021). The recent use of machine learning (ML) tools has accelerated the biomarker discovery process for psychiatric disorders. The ML tools have already provided gene expression markers with higher diagnostic potential (Hess et al., 2020; Liu et al., 2022; Zhu et al., 2021). Currently, the ML-based *in silico* approaches are limited to publicly available microarray datasets. ML-based *in silico* studies using biomarkers have reported higher diagnostic performance for psychiatric disorders (Ke et al., 2021; Wu et al., 2022; Yu et al., 2016; Zhu et al., 2021). However, few studies have validated their ML models using independent test datasets (Hess et al., 2020, 2016; Liu et al., 2022). Application of the diagnostic models into clinics would need extensive validation and appropriate data scaling methods to develop robust models. In addition, most of the studies focused on accuracy and area under the curve (AUC) as evaluation parameters for the performance of ML models. However, schizophrenia is associated with a higher degree of stigmatization and demands a diagnostic test with higher precision (True positives / (True positives + False positives)).

In this study, we have attempted to develop an ensemble of ML algorithms to classify SCZ samples. We selected publicly available gene expression microarray datasets for this meta-analysis. The raw data from each platform was processed independently to avoid data leakage. ML models were developed using support vector machines (SVM) and prediction analysis for microarrays (PAM) algorithms. We made use of differential gene expression analysis (DGEA) for selecting features with potential diagnostic values. ML models with different sets of genes were compared based on their performance in test data class prediction. The test data predictions from best-performing models were ensembled to achieve higher precision. The gene ontology and networking analysis of the feature genes further highlighted the biological processes and hub genes associated with SCZ. These ensemble models were finally tested for cross-platform compatibility.

## 2. Materials and methods

### 2.1. Identification of datasets

Peripheral blood gene expression microarray datasets for schizophrenia (SCZ) were identified from the Gene Expression Omnibus (GEO) (Edgar et al., 2002) and ArrayExpress (Parkinson et al., 2007) using keywords ‘Gene expression’, ‘Peripheral blood’, ‘Biomarkers’ and ‘Schizophrenia’ or ‘Schizophrenia spectrum’. A similar search was performed on databases such as PubMed and Google Scholar. Studies with immortalized cell lines, specific cell types, and custom microarray platforms were excluded. The analysis included studies with available raw data, while the authors were contacted to obtain the data for studies where it was not publicly available.

### 2.2. Importing and processing of raw data

Raw data for each dataset was imported and processed independently in R (R Core Team, 2020). Probe filtration was carried out for Illumina datasets (e.g. Illumina probes with detection P.val <0.05 in _≥_ 3 samples were retained). Probe IDs of all the arrays were mapped to HUGO Gene Nomenclature Committee (HGNC) gene symbols (Braschi et al., 2019). Gene expression values for multiple probes were averaged out for individual genes. All the datasets were combined based on the common genes to obtain a meta-file. This meta-file with raw gene expression values (raw meta-file) was processed for the identification of outlier datasets and for machine learning (ML) based prediction analysis.

### 2.3. Identification of outlier datasets

We made use of the expression status of differentially expressed genes for the identification of outlier datasets. Before differential gene expression analysis, samples from raw meta-file were independently quantile normalized based on the microarray platform used. Illumina, single-channel Agilent, and Affymetrix datasets were normalized using lumi (Du et al., 2008), limma (Ritchie et al., 2015), and affy (Gautier et al., 2004) packages, respectively. The normalized data was further batch corrected using ComBat (Leek JT, Johnson WE, Parker HS, Fertig EJ, Jaffe AE, Zhang Y, Storey JD, 2020) and subjected to differential gene expression analysis (DGEA) using limma. The expression status of top differentially expressed genes was visualized for heterogeneity in their expression across the individual datasets using a Forest-plot (Gordon and Lumley, 2021).

### 2.4. Pre-processing and data scaling for machine learning

To avoid any data leakage, the raw meta-file (raw gene expression values) was divided into train and test data before normalization and batch correction. To achieve this, samples were shuffled and then subjected to a random selection of train (90%) and test (10%) data (Figure 1A). This random selection was repeated to obtain 10 iterations of train and test datasets. Samples within the training data were quantile normalized based on the microarray platform (Figure 1B). Training data was further batch-corrected independently (Figure 1B). In contrast, test data was normalized using quantile targets (Bolstad, 2020) from train data and batch corrected using train data as reference (Figure 1B) (Leek JT, Johnson WE, Parker HS, Fertig EJ, Jaffe AE, Zhang Y, Storey JD, 2020). Each iteration of normalized and batch-corrected train data was used for feature selection and development of ML models. In contrast, test data was used to evaluate ML models (Figure 1C).

**Figure 1:**
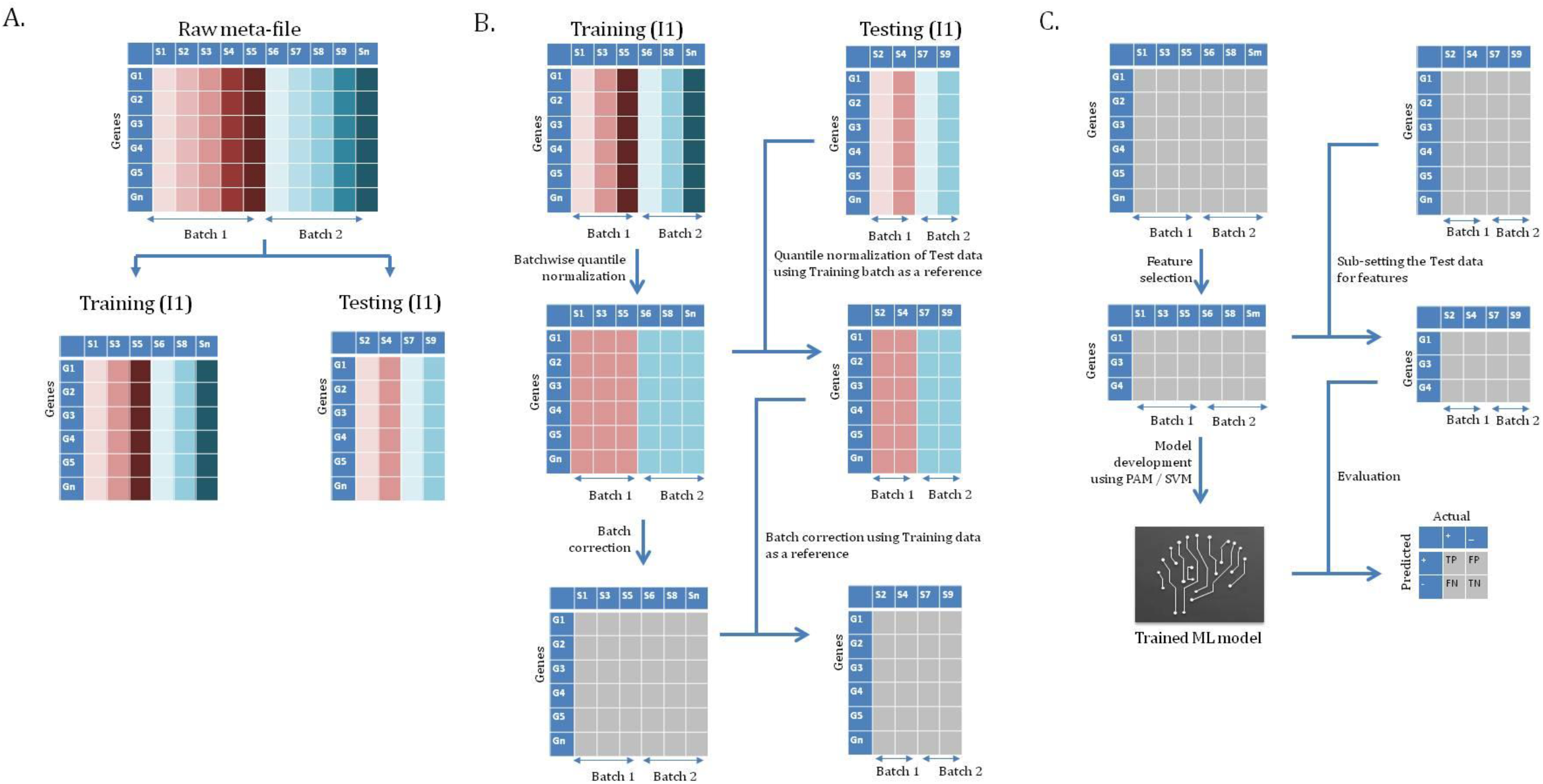
The workflow depicts the steps involved in sample processing and class prediction analysis. A) A raw meta-file with 6775 common genes (rows) across all the datasets and 449 samples (columns) was used for the analysis. Samples from the raw meta-file were shuffled and divided into train and test data (9:1). Random selection of was repeated to produce ten iterations of train and test data. B) Each iteration of train data was independently quantile normalized. The quantile targets for each batch in the train data were used to normalize respective test data. Normalized train data was batch corrected independently, while test data was batch corrected in reference to the respective train data. C) The pre-processed train data was used for feature selection and ML model. The pre-processed test data was used for model testing.

### 2.5. Feature selection and development of ML models

We used differential gene expression analysis (DGEA) as a feature selection method. The top differentially expressed genes were used as features for model building. Support vector machines (SVM) (David Meyer et al., 2021) with different kernels (linear, polynomial, radial and sigmoid) and prediction analysis for microarrays (PAM) (Hastie et al., 2019) algorithms were used for the development of diagnostic models for SCZ. The ML models built from each iteration of the train data was cross-validated (k=10) and were used for class prediction of respective test data samples (Figure 1C). The ML models were evaluated based on accuracies and were selected for ensemble learning.

### 2.6. Ensemble learning and evaluation

In order to improve the precision of class prediction, we used ensemble learning of ML models of SVM and PAM. In brief, samples predicted as cases by both ML algorithms were labelled as cases in ensemble models. The performance of the ensemble model was evaluated in comparison with individual models using parameters such as precision, accuracy, sensitivity and specificity.

### 2.7. Functional enrichment and networking analysis

Functional enrichment analysis for the genes of interest was carried out using g:profiler. A search tool for the retrieval of interacting genes/proteins (STRING) (Szklarczyk et al., 2019) based protein-protein interaction network (PPI) for these genes was established in Cytoscape (Shannon et al., 2003). The essential nodes (hub genes) of the PPI network were identified by the maximal clique centrality (MCC) method of the cytohubba plugin (Chin et al., 2014). A network of hub genes and their first-stage nodes were later visualized in Cytoscape.

### 2.8. Establishment of case-control study

Protocol for this study was approved by KEM Hospital Research Centre Ethics Committee (KEMHRC ID No. 2001) and Symbiosis International (Deemed University) Independent Ethics Committee (SIU/IEC/99). We recruited 20 participants of the age group 18-65 years from the Psychiatry Unit K.E.M hospital, Pune. Consent from all the participants was obtained before recruitment. Consent for participants with suspected case of SCZ was supported by the consent of first-degree relatives.

The common exclusion criteria for control (CNT) and schizophrenia (SCZ) groups were the presence of a) acute or chronic infections, b) coronary heart disease, c) metabolic disorders, d) arrhythmia, e) heart disorders, f) hyper and hypothyroidism, g) inflammatory bowel disease and h) multiple sclerosis. Female participants with polycystic ovary syndrome, pregnant and lactating mothers, and women on in-vitro fertilization (IVF) treatment at the time of recruitment were also excluded from the study. The participants with a suspected case of schizophrenia and schizophrenia spectrum disorder were considered for recruitment under the SCZ group. Age and gender-matched participants with no history of psychiatric disorders were considered for recruitment under the control (CNT) group. All the participants, irrespective of the group, were subjected to diagnosis.

### 2.9. Clinical interview and diagnosis

SCZ diagnosis was carried out using structured clinical interview for DSM-5 research version (SCID-5-RV) (version 1.0.0) (First MB, Williams JB, Karg RS, 2015). The SCID-5-RV was administered by a trained psychiatrist and a psychologist. The SCZ diagnosis was later confirmed by a senior psychiatrist from the team. SCZ-diagnosed participants were also administered with positive and negative syndrome scale (PANSS) (Kay et al., 1987). The absence of any psychiatric disorder in the control group participants was confirmed by administering SCID-5-RV. Age, gender, family history for psychiatric disorders, medical history, and medication status were recorded for all the participants.

### 2.10. Blood collection and RNA extraction

Random (non-fasting) venous blood samples were collected in K2EDTA vacutainers and processed on the same recruitment day. A blood cell count (hemogram) was performed on the samples collected. Peripheral blood mononuclear cells (PBMCs) were isolated using Ficoll-Paque (Sigma, Catalogue: GE17-5442-02) density gradient centrifugation and re-suspended in TRIZOL (ThermoFischer Scientific, Catalogue: 15596026). Samples were subjected to RNA-sequencing (RNA-Seq) using commercial services. In brief, ribo-depleted RNA samples were sequenced using NovaSeq 6000 system - Illumina to obtain a minimum of 60 million paired-end reads of 150 nucleotide length.

### 2.11. Pre-processing of RNA-Sequencing data

The quality of each sample was confirmed using FastQC (Andrews, 2010). The sequences were aligned to the human genome (GENECODE hg38) (Frankish et al., 2019) using HISAT2 (Kim et al., 2019). The aligned files were subjected to gene assignment using featureCounts (Liao et al., 2014) to create a count matrix. The gene expression values were locally normalized by converting the raw count to counts per million (CPM) and transcript per million (TPM). The raw counts (RC), CPM and TPM matrix were further quantile normalized and batch corrected in reference to microarray training datasets independently. The quantile normalized and batch-corrected RC, CPM, and TPM matrices were further used to evaluate the cross-platform performance of ML models developed using microarray data.

### 2.12. Statistical analysis

Data from processed microarray datasets and patient samples were analyzed using Microsoft Excel-Real Statistics (Zaiontz, 2020) and PAST (Hammer et al., 2001). Microarray datasets processed using PAM, SVM and ensemble approach were tested for normality using the Shapiro-Wilk test. For data with normal distribution, one-way ANOVA with an alpha of 0.05 followed by Tukey’s post hoc test with Dunn–Šidák correction was performed. For clinical samples, age and blood cell count data were tested for normality as mentioned previously. The difference between control and case groups was studied using an unpaired t-test for normal datasets and Mann-Whitney U test for non-normal data. The gender for control and case groups were compared using Chi-square test respectively.

## 3. Results

### 3.1. Microarray datasets show variability in gene expression with no outliers

We identified seven peripheral blood expression array datasets for SCZ (Table 1). Participants from these datasets belonged to different ethnic groups. Most of the datasets identified had medicated or a mixed population of SCZ participants. For Kumarasinghe et al., paired study, only the ‘before treatment’ samples were considered for the analysis to avoid over-representation of the same samples. Only the genes common to all seven datasets were retained for the meta-analysis. The resulting metafile contained 449 samples with 6775 genes.

**Table 1:** Schizophrenia Microarray gene expression datasets

| Dataset | Platform | Control /SCZ | Female (%) | Medication status | Genes analysed | Ethnicity or origin |
| --- | --- | --- | --- | --- | --- | --- |
| GSE18312 (Bousman et al., 2010) | Affymetrix Human Exon 1.0 ST Array | 8+13 | 33 | Medicated | 17131 | San Diego and Taiwan |
| GSE27383 (van Beveren et al., 2012) | Affymetrix Human Genome U133 Plus 2.0 Array | 29+43 | NA | Mix | 21826 | Multi ethnic groups |
| GSE38481 (de Jong et al., 2012) | Illumina HumanRef-8_V3 beadchip | 22+15 | 27 | Mix | 12647 | Denmark and Netherland |
| GSE38484 (de Jong et al., 2012) | Illumina HumanHT-12_V3 beadchip | 96+106 | 42 | Mix | 17233 | Denmark and Netherlands |
| GSE48072 (Stoll et al., 2013) | Illumina HumanHT-12_V4 beadchip | 31+35 | 53 | NA | 15155 | Finland, Sweden, Caucasian |
| GSE54913 (Xu et al., 2016) | Arraystar Human LncRNA microarray V2.0 | 12+18 | NA | Treatment naive | 13003 | Han Chinese |
| Kumarasinghe et al. 2013 (Kumarasinghe et al., 2013) | Illumina HumanHT-12_V3 beadchip | 11+10 | 38 | Treatment naive | 10544 | Sinhalese |

**Note:** The “Mix” medication status indicates that the participants with and without drug treatment were part of the study. While, “NA” indicates unavailability of information on medication status.

The differential gene expression analysis (DGEA) of the quantile normalized and batch-corrected meta-file (Supplementary figure 1) resulted in 1988 DEGs in SCZ samples with respect to controls (adj. P val <0.05) (Supplementary figure 2). Heterogeneity among the datasets was observed by Forest-plot of two up-regulated (*CLEC5A* and *EIF1AY*; LFC >1) (Supplementary figure 3A and 3B) and two down-regulated (*EOMES* and *EHMT2*; LFC <-1) DEGs (Supplementary figure 3C and 3D). The expression pattern of the selected DEGs varied across all datasets with respect to the mean expression status; however no specific dataset could be identified as an outlier. Hence, all the datasets were retained for the analysis.

### 3.2. Ensemble learning results in higher precision for schizophrenia diagnosis

The pre-processing of raw meta-file resulted in normalized (Supplementary figure 4) and batch-corrected (Supplementary figure 5) train and test datasets. The DGEA of each train data iteration resulted in the identification of DEGs as features (Supplementary figure 6). Machine learning models were built using these feature genes from training datasets.

We used two different ML models, support vector machines (SVM) and prediction analysis for microarrays (PAM), to classify test data samples. The performance of these ML models was evaluated based on the mean test data prediction accuracy. SVM models with kernels such as “linear”, “polynomial”, “radial”, and “sigmoid” kernels did not show any significant difference in the test data prediction accuracy (data not shown). We chose SVM-radial for further analysis. A comparison of SVM-radial models with different numbers of features (top5, top25, top100, top400, top1600, and all genes) revealed that the performance of SVM-radial drops with features more than 400 DEGs (Figure 2A). PAM models with different numbers of features did not impact the test data prediction accuracy (Figure 2B). SVM-radial and PAM models were further used for combinatorial analysis.

**Figure 2:**
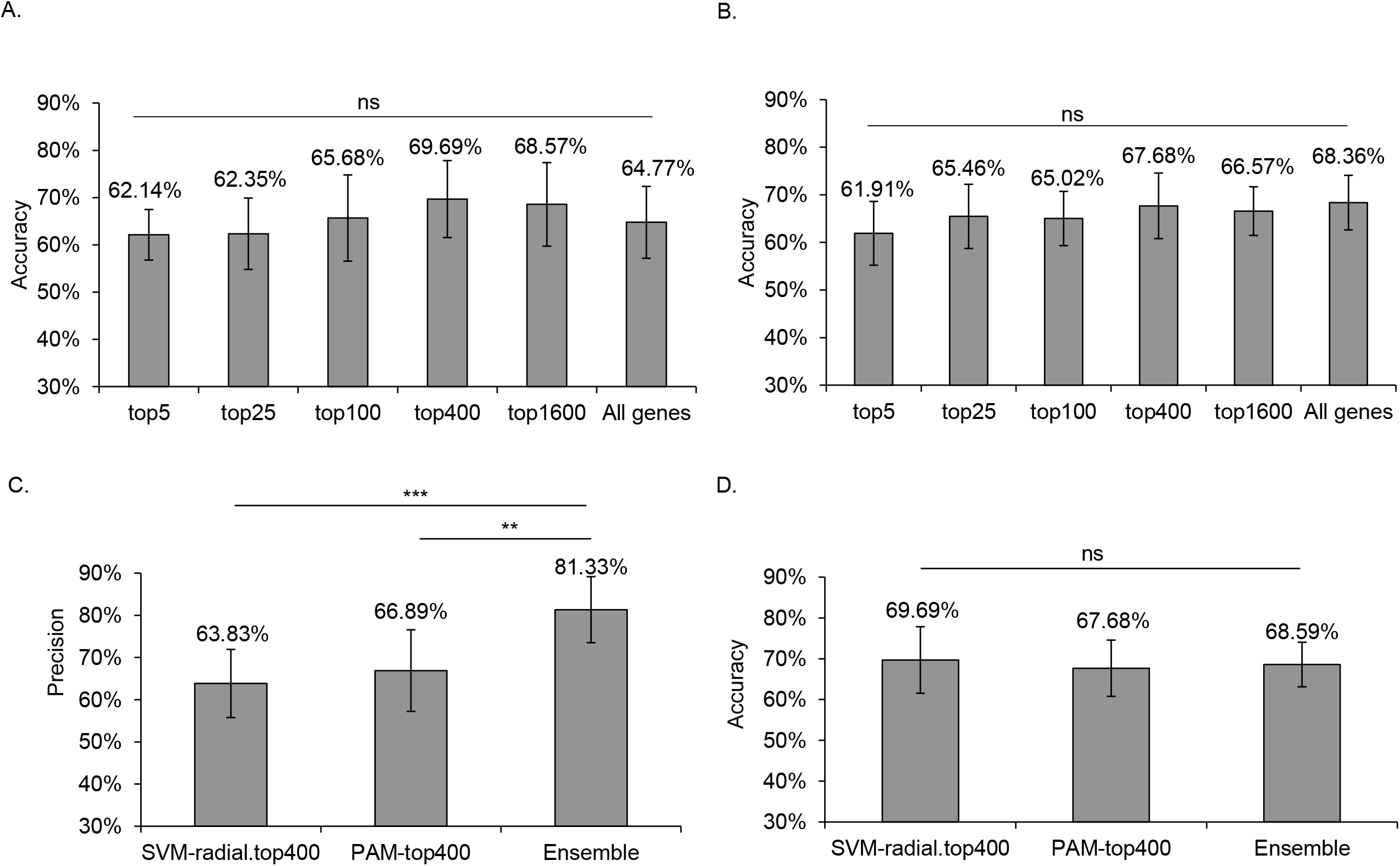
Test data prediction accuracy and precision of SVM-radial, PAM and ensemble models with differentially expressed genes (DEGs) as features. Machine learning models with different number of features were compared using one-way ANOVA followed by Tukey’s post hoc test with Dunn– Šidák correction. A) No significant difference was observed between SVM-radial models with different number of features. B) Similar comparison of PAM models also resulted in no significant difference. The ensemble of SVM-radial and PAM models with top 400 DEGs outperformed the individual models. C) The test data classification precision for ensemble model with top 400 DEGs was significantly high compared to the SVM-radial and PAM. D) The drop in the accuracy of ensemble model was not significant.

We ensemble the select ML models of SVM-radial and PAM for this combinatorial analysis. In brief, only the samples identified as cases by both algorithms were classified as cases to reduce false positives in test data class prediction. The ensemble of SVM-radial and PAM with top 400 DEGs (ensemble-400) had the highest precision of 81.33% (SD: 0.078) compared to all other ensemble models (Supplementary figure 7A). Also, the accuracy of ensemble-400 (68.59%, SD: 0.055) and ensemble-all genes (69.48%, SD: 0.048) was significantly high compared to ensemble-5 (60.37%, SD: 0.057) (Supplementary figure 7B). However, no significant difference was observed in the sensitivity and specificity of the ensemble models (Supplementary figure 7C and 7D). We chose ensemble-400 based on the absolute value (expressed in percentage) of precision for further analysis. The precision of ensemble-400 was significantly higher compared to the SVM-radial: 66.83 (SD: 0.081) and PAM: 66.89% (SD: 0.097) (Figure 2C). The ensemble learning achieved higher precision without a significant drop in accuracy when compared to individual models (Figure 2D). Interestingly, the decrease in sensitivity of ensemble-400 was not significant when compared to SVM-radial and PAM (Supplementary figure 8A and 8B).

### 3.3. Functional enrichment and network analysis identify biological processes, pathways and hub genes associated with schizophrenia

The top 400 DEGs as features in combinatorial analysis predicted SCZ samples with the highest precision (lesser false positives). We identified common genes (n: 207) between the top 400 DEGs from all ten iterations of training datasets. Functional enrichment analysis of the common genes identified apoptosis and natural killer cell-mediated cytotoxicity as the top two enriched Kyoto encyclopaedia of genes and genomes (KEGG) pathways (adj.P.val <0.05). The majority of DEGs were enriched for the biological processes associated with immune function. However, the top biological process enriched were response to stimulus and response to stress (Figure 3A). Metabolic processes such as the organonitrogen compound metabolic process and regulation of nitrogen compound metabolic process were also significantly enriched. We identified the top 5 hub genes (*PRF1, GZMB, IL2RB, ITGAL*, and *IL2RG*) from the PPI network of common genes (Figure 3B).

**Figure 3:**
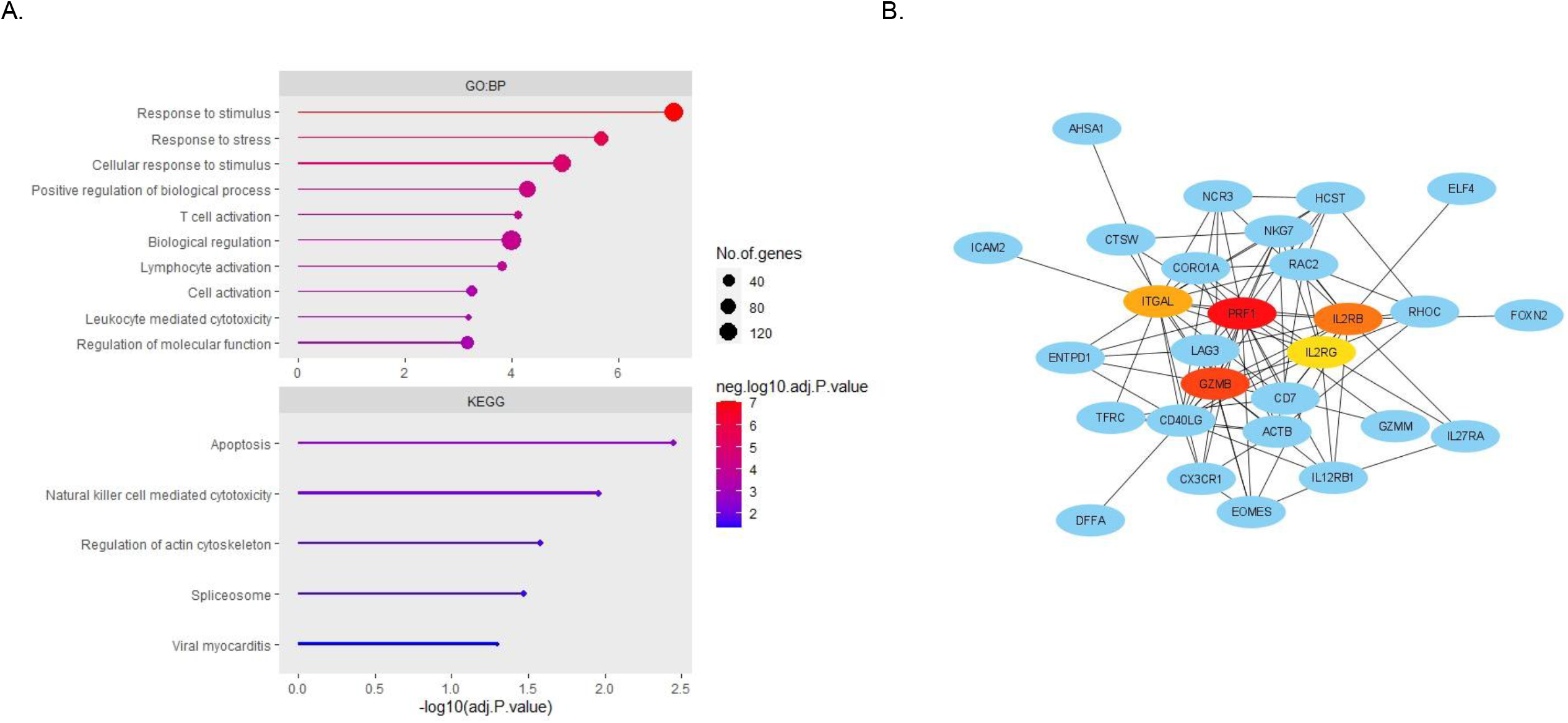
Functional enrichment and networking analysis was performed using the common genes from top 400 DEGs of ten iterations of training sets. The enriched top 10 biological processes (adj.P.val <0.05) and KEGG pathways were visualized using a bar-plot. A STRING based protein-protein network was established for common genes using Cytoscape. The maximal clique centrality (MCC) method of cytohubba plugin identified top 5 hub genes. The network of 5 hub genes and their first-degree nodes was visualized. The intensity of node colour represents the MCC rank. i.e., PRF1 ranked as number one followed by GZMB, IL2RB, ITGAL, and IL2RG respectively.

### 3.4. External cross-platform validation of ensemble models

A case-control study was established with 20 participants of Indian origin (SCZ: 10, CNT: 10). There was no significant difference in age, gender, and blood cell counts between case and control group participants (Table 2). All the SCZ-affected participants recruited in this study were on antipsychotic medication. Ribonucleic acid (RNA) isolated from the peripheral blood samples of the participants were subjected to RNA-Sequencing analysis. The sequencing resulted in ∼80 million reads per sample. The quality of sequencing was satisfactory (Supplementary figure 9) which resulted in acceptalble percent alignment (Supplementary table 1). The RNA-Sequencing data was used for external cross-platform validation of the models. The class prediction analysis was performed using raw counts (RC), counts per million (CPM) and transcript per million (TPM) matrices, as mentioned in the methodology section. The prediction accuracy of ML models with TPM counts was relatively better compared to CPM and RC (data not shown). ML models’ performance with TPM counts is reported in this analysis. SVM-radial (51.50%, SD: 0.05), PAM (58.50%, SD: 0.02) and ensemble with top 400 DEGs (62.50%, SD: 0.03) predicted RNA-Seq test samples with low accuracy (Figure 4A). However, the ensemble-400 was able to classify SCZ samples with moderate precision (72.00%, SD: 0.08), which was significantly higher than PAM (57.00%, SD: 0.05) and SVM-radial (47.00%, SD: 0.11) (Figure 4B). The higher precision of ensemble models was accompanied by higher specificity and lower sensitivity compared to the individual models (data not shown).

**Table 2:** Demographics and selected features of cell types in case-control study

| Characteristics | SCZ | Control | P value |
| --- | --- | --- | --- |
| Age (Year)* (n: 20) | 33.0 (9.754) | 36.6 (6.04) | 0.4988 |
| Gender (M/F)# (n: 20) | 3/5 | 4/6 | 0.9200 |
| Neutrophil (103/μL)# | 4.8 (2.04) | 3.9 (1.26) | 0.2810 |
| Lymphocyte (103/μL)# | 2.3 (0.68) | 2.2 (0.43) | 0.8623 |
| Monocyte (103/μL)# | 0.5 (0.13) | 0.6 (0.11) | 0.3617 |
| Eosinophil (103/μL)\$ | 0.2 (0-0.3)* | 0.2 (0.1-0.7)* | 0.2369 |
| Basophil (103/μL)\$ | 0.025 (0-0.1)* | 0.040 (0-0.1)* | 0.9650 |

**Note:** SCZ: Schizophrenia, CNT: Control, \* Chi-squared test, # Unpaired t-test (s.d.), \$ Mann-Whitney U test (min-max). Note: Since cell counts for two samples were not available the comparison of blood cell counts between SCZ and CNT was performed on 18 samples.

**Figure 4:**
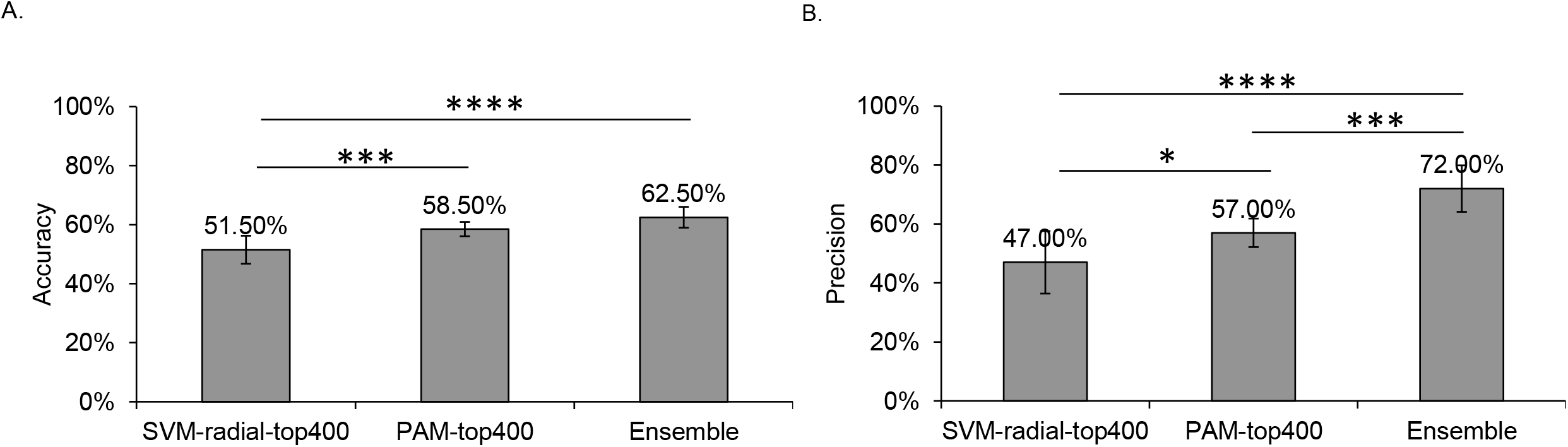
Cross-platform validation of SVM-radial, PAM and ensemble models with top 400 differentially expressed genes. Microarray based ensemble learning was tested for their ability to classify schizophrenia samples from RNA-Sequencing platform. A comparative analysis between the models was performed using one-way ANOVA followed by Tukey’s post hoc test with Dunn– Šidák correction. A) Ensemble models classified test samples with significantly higher accuracy when compared to individual models. B) Similarly, ensemble models classified test samples with significantly higher precision compared to the individual models.

## 4. Discussion

A neuropsychiatric disorder such as schizophrenia is associated with stigmatization. Hence, a diagnostic test with higher precision is desirable. In this study, we attempted to develop an ensemble model with higher precision for classifying SCZ samples. The ensemble model resulted in the test data precision of 81.33% (SD: 0.078), without any significant drop in accuracy. We developed a unique strategy of pre-processing microarray data to build a robust model that can be applied to a single sample level for clinical application.

Microarray data was pre-processed for building ML models. Seven different datasets with participants from varied ethnicity were included in this analysis. The pre-processing of data involved shuffling of samples, division of samples into train and test data, normalization, and batch correction. Shuffling of samples ensured the representation of each dataset in train and test data. The ten-fold split of samples into train and test data before normalization prevented data leakage. The train data was pre-processed independently, whereas test data was normalized and batch-corrected using train data as reference (Figure 1B). This pre-processing of test data removed the existing technical differences and allowed us to independently process and predict each test sample. The pre-processing resulted in the development of robust models with a better chance of survival in clinics.

We used differential gene expression analysis (DGEA) for feature selection, which can be important for the application of models in clinical settings. Feature selection methods perform better than extraction methods for their “explainability” in clinical settings (Bhandari et al., 2022). In addition, simple filter methods such as DGEA are computationally less intensive as compared to embedded methods of feature selection. The comparison of ML models with different sets of DEGs revealed that the number of features does not affect the performance of PAM models, unlike SVM, for inexplicable reasons (Figure 2A and 2B). Further, we combined the test data predictions from SVM and PAM models to develop ensemble learning for higher precision. Both ML algorithms use different logic for classification. Thus, a consensus between the models resulted in higher precision without any significant drop in accuracy compared to the individual models (Figure 2C and 2D). The increase in precision was associated with increased specificity and an expected drop in sensitivity (Supplementary figure 9A and 9B). Each individual dataset used in this analysis showed a variation in prediction accuracy when tested using an ensemble model. The unavailability of medication status for each sample did not allow us to assess the impact of medication status on test data prediction. It should be noted that the antipsychotic-treated samples from GSE18312 showed an intermediate accuracy when compared to the drug-naive samples from Kumarasinghe et al. and GSE54913 datasets. Similarly, the mixed medication status samples from GSE27383, GSE38481, and GSE38484 did not show a variation in overall accuracy or precision. Precision on the higher side of 60% for each dataset suggested our prediction algorithm’s robustness (Supplementary figure 10).

We could identify very few studies using ML models for SCZ class predictions. *In silico* analysis by Jonathan Hess et al., reported an area under the curve (AUC) of 0.72 to 0.77 for SCZ Vs CNT and 0.607 for bipolar disorder (BD) Vs SCZ in two separate studies with independent test datasets (Hess et al., 2020, 2016). A recent study reported a much higher AUC of 0.993 with 10-fold cross-validation for the classification of SCZ samples from that of controls (Zhu et al., 2021). However, the higher performance of the models in this study can be attributed to the uniform population with fewer confounding factors such as ethnicity. We did not come across a study with the aim of developing ML models with higher precision using ensemble learning. The previous studies with a multi-modal approach suggest the need to integrate biological and clinical information for better performance of ML models (Fernandes et al., 2020; Ke et al., 2021). However, the unavailability of clinical information for each sample in the publicly available GEO datasets restricted this analysis to only gene expression markers.

Ensemble of SVM-radial and PAM with top 400 DEGs (Ensemble-400) classified test data samples with the highest precision (Figure 2C). We identified common genes (n: 207) between the top 400 DEGs in the ten iterations of training datasets. Enrichment analysis of these common genes highlighted key pathways such as apoptosis and natural killer cell-mediated cytotoxicity (Figure 3A), which have been known to be dysregulated in SCZ (Parellada and Gassó, 2021; Yovel et al., 2000). The biological processes related to immune function and organonitrogen compound metabolic process have also been associated with SCZ earlier (Dmitrieva et al., 2022; Van Kesteren et al., 2017). In addition to biological processes and pathways, we also identified the key regulators (hub genes) of the protein-protein interaction network (Figure 3B). These hub genes were involved in top enriched biological processes such as response to stress and response to stimulus. The hub genes identified in this study have been previously reported in association with neuropsychiatric disorders, including SCZ (Fallin et al., 2005; Ghazaryan et al., 2014; Ibrahim et al., 2017). The genome wide association studies have also associated *PRF1, GZMB*, and *IL2RB* with SCZ (Pardiñas et al., 2018; Ripke et al., 2014). Further, *IL2RB, ITGAL* and *IL2RG* are known to be differentially expressed in the peripheral blood of SCZ-affected individuals (Ghazaryan et al., 2014; Leirer et al., 2019). Interestingly, only *ITGAL* and *IL2RB* have been reported to be differentially expressed in the first episode SCZ affected individuals (Leirer et al., 2019). Of these 207 DEGs, *MAP4K1, GOT2, MCM3, SIGIRR, SRPK1, TIPARP, RPRD1A, ATIC, NKG7*, and *SCAP* were also highlighted in our previous study for their association with SCZ (Wagh et al., 2021)

We also performed cross-platform validation of the ensemble model using RNA-Sequencing data. These samples were not part of the machine learning model development and hence served as external test data for the validation of the models. To achieve this, we established a case-control study with age and gender-matched participants. The validation study included pre-processing of RNA-Sequencing data to generate transcript per million (TPM), counts per million (CPM) and raw counts. These values were batch-corrected in reference to microarray train data and the samples were predicted by ML models independently. The predicted accuracy of test data with TPM values was relatively higher compared to CPM and RC (data not shown) suggesting the compatibility of TPM values for cross-platform validation studies. Similar to microarray data, the ensemble model predicted SCZ samples with significantly better precision as compared to individual models (Figure 4B). Prediction analysis for microarrays (PAM) models performed significantly better when compared to the support vector machines (SVM-radial) in cross-platform data prediction. However, overall low accuracies of individual models suggest a need for the development of better cross-platform normalization techniques (Figure 4A).

## 5. Strengths and limitations of the study

To the best of our knowledge, this is the first study that uses ensemble learning for schizophrenia (SCZ) diagnosis with higher precision. The higher precision offered by ensemble learning, even with the existing diversity in the samples with respect to ethnicity, age, gender, and medication status, indicates the robustness of the models. The pre-processing of raw microarray data in this analysis ensures no data leakage and allows the prediction of a single test sample. Notably, cross-platform validation confirms the compatibility of transcript-per-million (TPM) normalization of RNA-Sequencing data with microarray-based machine learning (ML) models for prediction analysis. The pipeline established in this study is not limited to SCZ and can be used for any disorder associated with a higher degree of stigmatization. There are several limitations of this study. The unavailability of clinical information for each sample restricted its use in developing multi-modal ML models. The performance of ML models in cross-platform validation was relatively poor, suggesting scope for developing better normalization methods. The analysis was restricted to only SCZ and did not attempt multi-class classification as Yang et al. did in their study (Yang et al., 2022). The modest sample size of the case-control study established did not allow us to explore Indian scenario for schizophrenia in detail.

## 6. Conclusions and future directions

In conclusion, we provide proof of concept for developing robust predictive models with higher precision for diagnosing SCZ. The current strategy effectively deals with the problems like data leakage and pre-processing of single microarray samples. The feature genes and biological pathways identified in this study can be pursued to explore their potential role in the disorder. Most importantly, this study attempted cross-platform class prediction using RNA-Sequencing data as test samples. However, a relatively poor cross-platform performance indicates the need for better cross-platform normalization techniques. In addition, the availability of data from other high throughput genome-wide studies may create novel avenues for developing multi-modal learning. Specifically, the multi-omic approach integrating genomic, transcriptomic, and proteomic data will surely result in the precise diagnosis of psychiatric disorders.

## Data Availability

Data availability:
The RNA-Sequencing data of this study will be available from the corresponding authors upon publication.
Code availability:
The R scripts used for the analysis are available on GitHub (https://github.com/macdlab/2023_VW_SCZ_Ensemble)

## Declarations

### Compliance with ethical standards

Two independent ethical committees approved the study protocol, KEM Hospital Research Centre Ethics Committee (KEMHRC ID No. 2001) and Symbiosis International (Deemed University) Independent Ethics Committee (SIU/IEC/99). Informed consent was obtained from all the participants. The consent for participants with schizophrenia was supported by the consent of a first-degree relative. Clinical interviews were administered by a trained psychiatrist and a psychologist. The diagnosis was confirmed by a senior psychiatrist. All the participants were compensated for their travel and time.

## Author contributions

VVW performed sample processing and data analysis, and wrote the manuscript. SA, SP, and VP recruited participants, collected data, and contributed to writing the clinical aspects. TP carried out a statistical analysis. LN assisted in setting up the data analysis pipeline and contributed to writing the manuscript. SPK and VP designed the study and wrote the manuscript. All authors discussed the results and approved the final version of the manuscript.

## Acknowledgements

We sincerely thank all the participants, their parents, relatives, and caretakers for their time and generous participation in making this project possible. We want to thank Deepa Raut, a phlebotomist, for her valuable contribution to the blood collection process. We would also like to thank Paul Tooney (Associate professor, New Castle University, Australia) for sharing data on request. We appreciate the assistance of Tanvi Kottat in proofreading the manuscript.

## Data availability

The RNA-Sequencing data of this study will be available from the corresponding authors upon publication.

## Code availability

The R scripts used for the analysis are available on GitHub (https://github.com/macdlab/2023_VW_SCZ_Ensemble).

