## Supplementary Figures for "Ensemble learning for higher diagnostic precision in schizophrenia using peripheral blood gene expression profile"

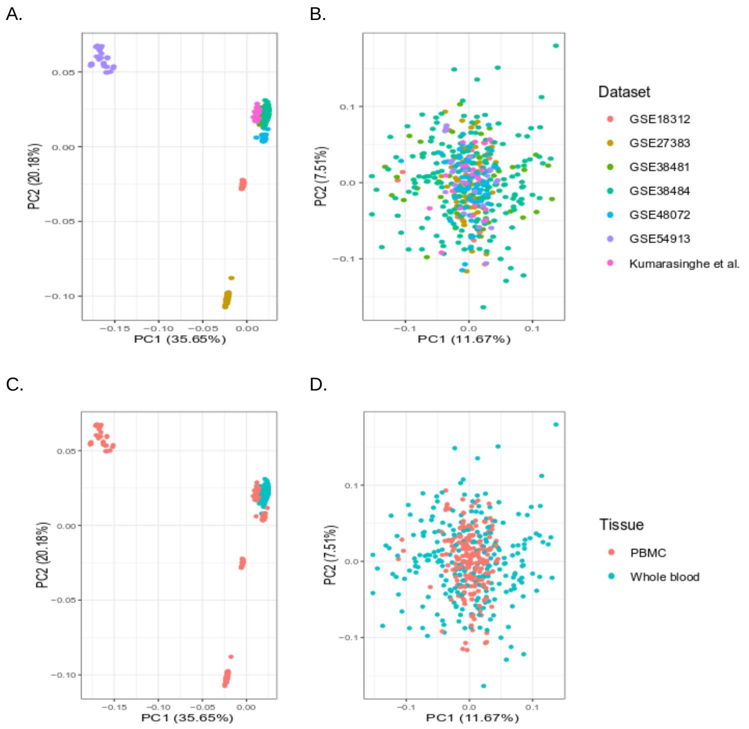


**Supplementary figure 1:** Principal component analysis (PCA) plots before and after batch correction. The raw-metafile was batch corrected for the microarray platform used in the datasets. A and B) The removal of batch effect was confirmed by visualizing distribution of samples across first two principal components (PC1 and PC2). C and D) The whole blood and PBMC dataset did not separate on either of the principal components.

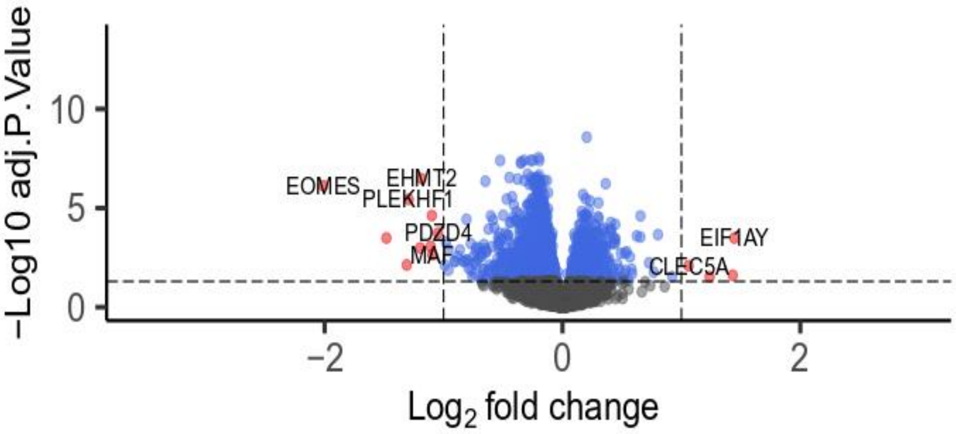


**Supplementary figure 2:** Volcano plot for normalized and batch corrected meta-file. The differential gene expression analysis using limma resulted in identification of top two up-regulated and top two down regulated genes. These genes were further used for identification of outlier datasets.

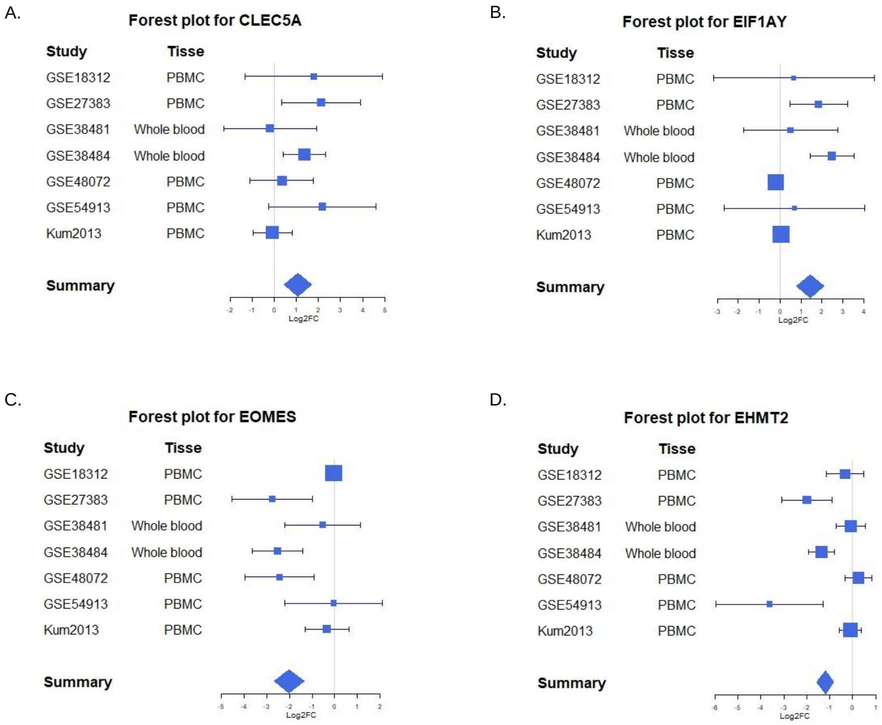


**Supplementary figure 3:** Forest plot for differential expression status of the top two up-regulated and top two down regulated genes across all the dataset. The expression status of DEGs from each dataset was compared to the mean expression of the DEGs in the batch corrected meta-file. The box size represents log FC values for each gene in the respective datasets and whiskers represents 95% confidence intervals. All the datasets were retained for the analysis since no specific trend in the expression status was observed irrespective of the tissue type (whole blood or PBMCs).

A.

B.

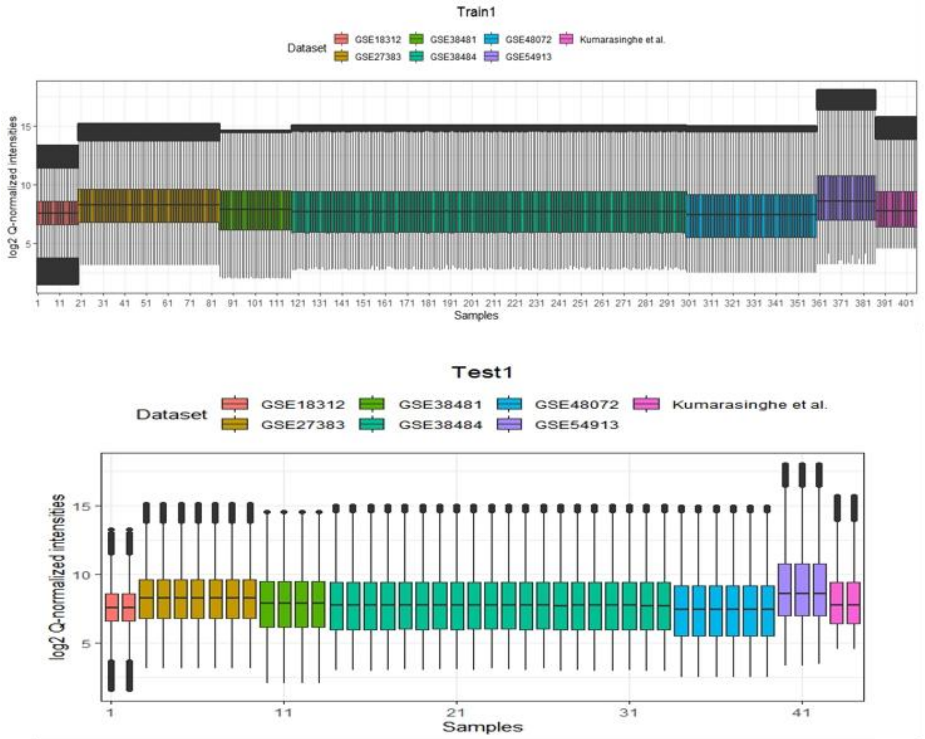


**Supplementary figure 4:** Boxplots for train and test data after quantile normalization. The pre-processing of raw meta-file for prediction analysis using machine learning models included quantile normalization of train and test datasets. A) The samples from each study in the train data was quantile normalized independently. B) The quantile targets from these studies were used for normalization of test samples from respective studies. The normalization was performed for each iteration of train and test datasets. The boxplots for iteration 1 was used as a representative image for visualization of quantile normalization.

A.

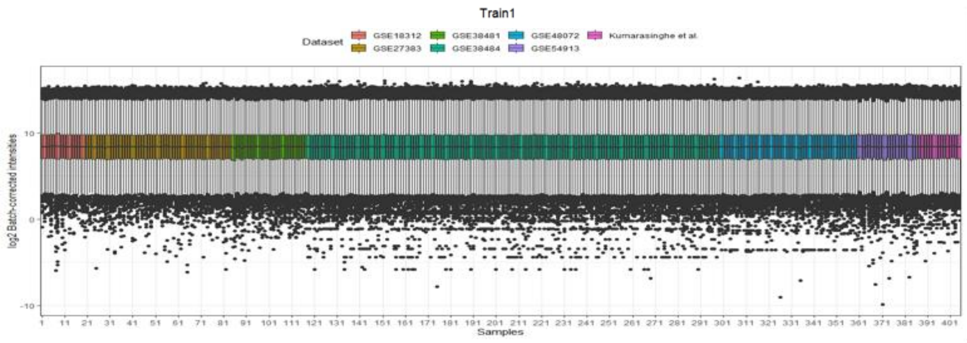


B.

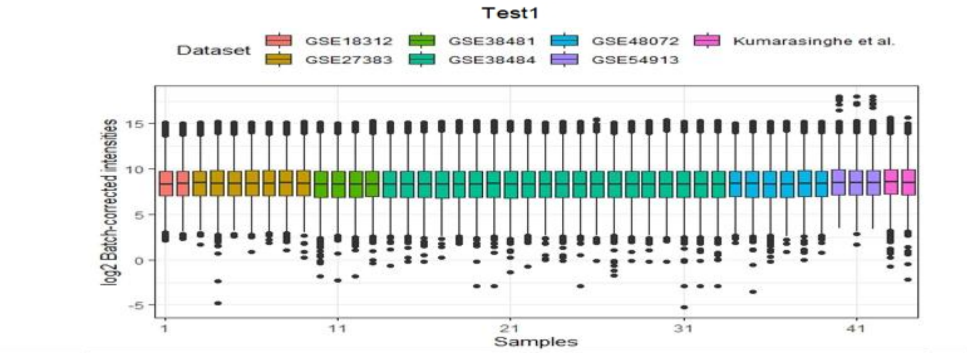


**Supplementary figure 5:** Boxplots for train and test data after batch correction. The quantile normalized train and test was batch corrected for the study types. A) Train data was batch corrected independently; B) whereas, test data was batch corrected using train data as reference. The boxplots for iteration 1 was used as a representative image for visualization of batch correction.

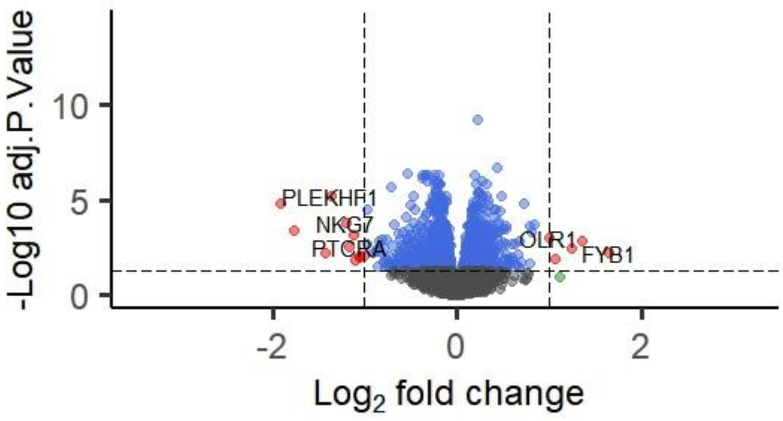


**Supplementary figure 6:** Volcano plot for normalized and batch corrected train data. The feature selection from pre-processed train data was performed using differential gene expression analysis (DGEA). The differentially expressed genes (adj.P.val < 0.05) were used as feature genes for development of ML models. The volcano plot for iteration 1 was used as a representative image for visualization of DGEA.

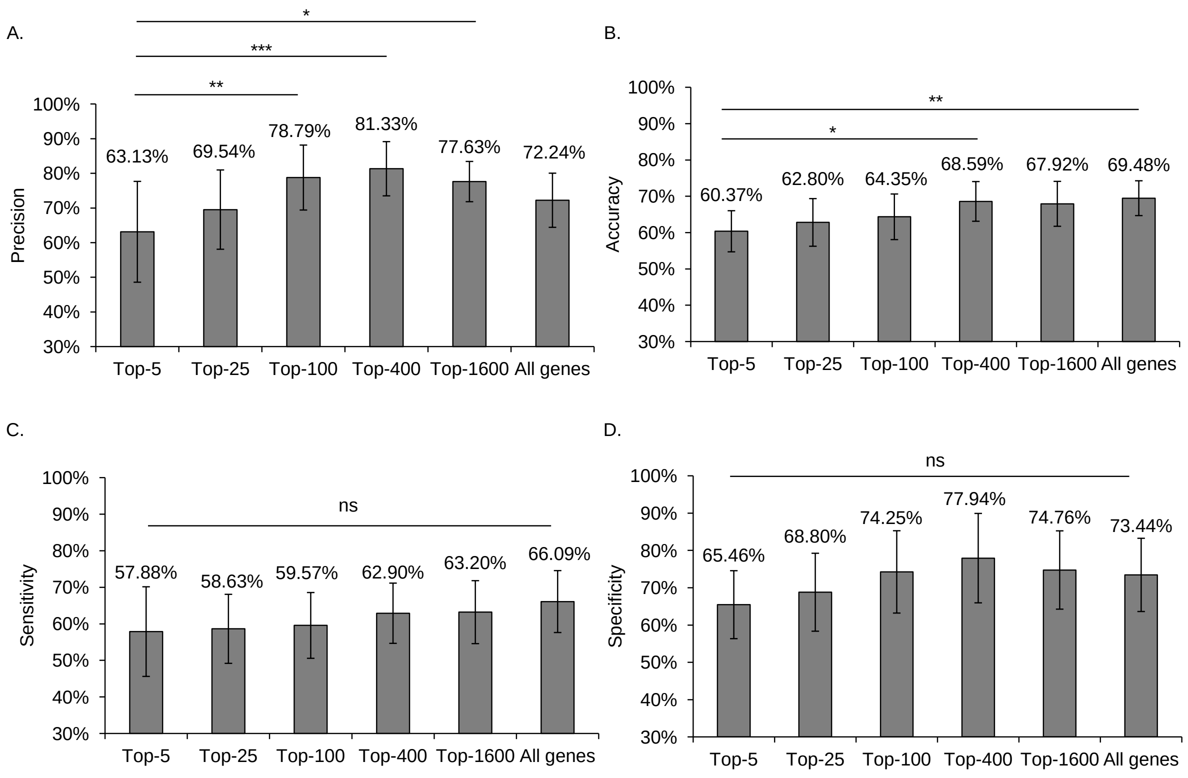


**Supplementary figure 7:** A comparative analysis of ensemble models in test data prediction. Ensemble models were compared using one-way ANOVA followed by Tukey's post hoc test with Dunn–Šidák correction. A) Precision of ensemble-100, ensemble-400, and ensemble-1600 was significantly higher than that of ensemble-5. Ensemble-400 had the highest precision compared to other ensemble models. B) Similarly, accuracy of ensemble-400 and ensemble-all genes was significantly higher than the ensemble-5. C and D) No significant difference between sensitivity and specificity of the ensemble models was observed.

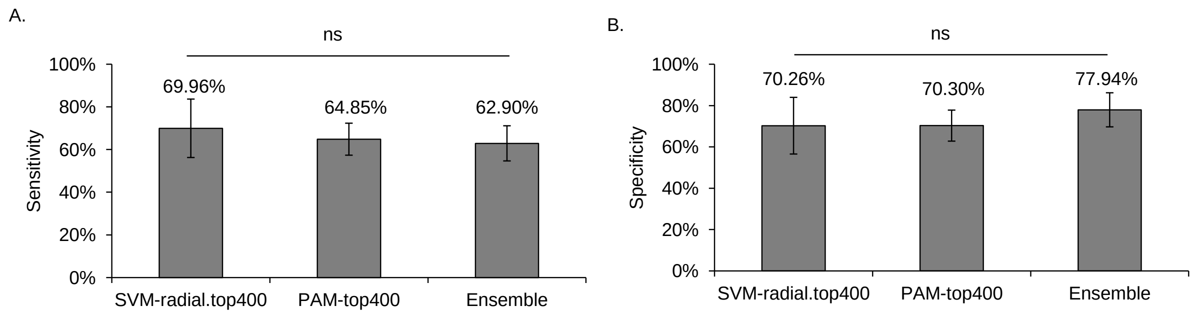


**Supplementary figure 8**: Test data prediction sensitivity and specificity of SVM-radial, PAM and ensemble model with top 400 differentially expressed genes. A and B) The one-way ANOVA followed by Tukey's post hoc test with Dunn–Šidák correction identified no significant difference in sensitivity and specificity of the models.

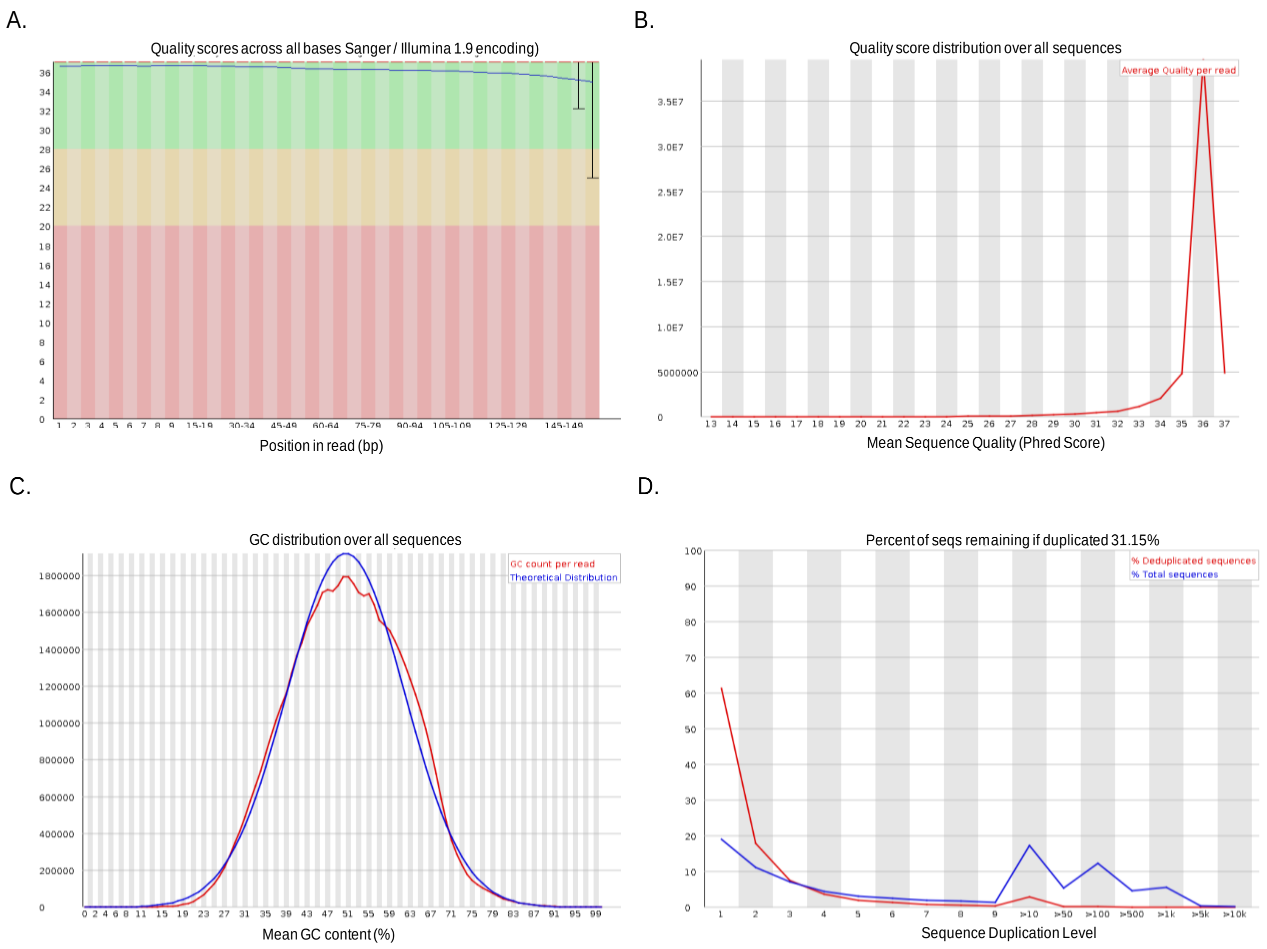


**Supplementary figure 9:** A representative FastQC report of an RNA-Seq sample. A) and B) suggest that the quality of sequencing was acceptable. B) and C) suggest no significant library contamination or PCR duplication.

A.

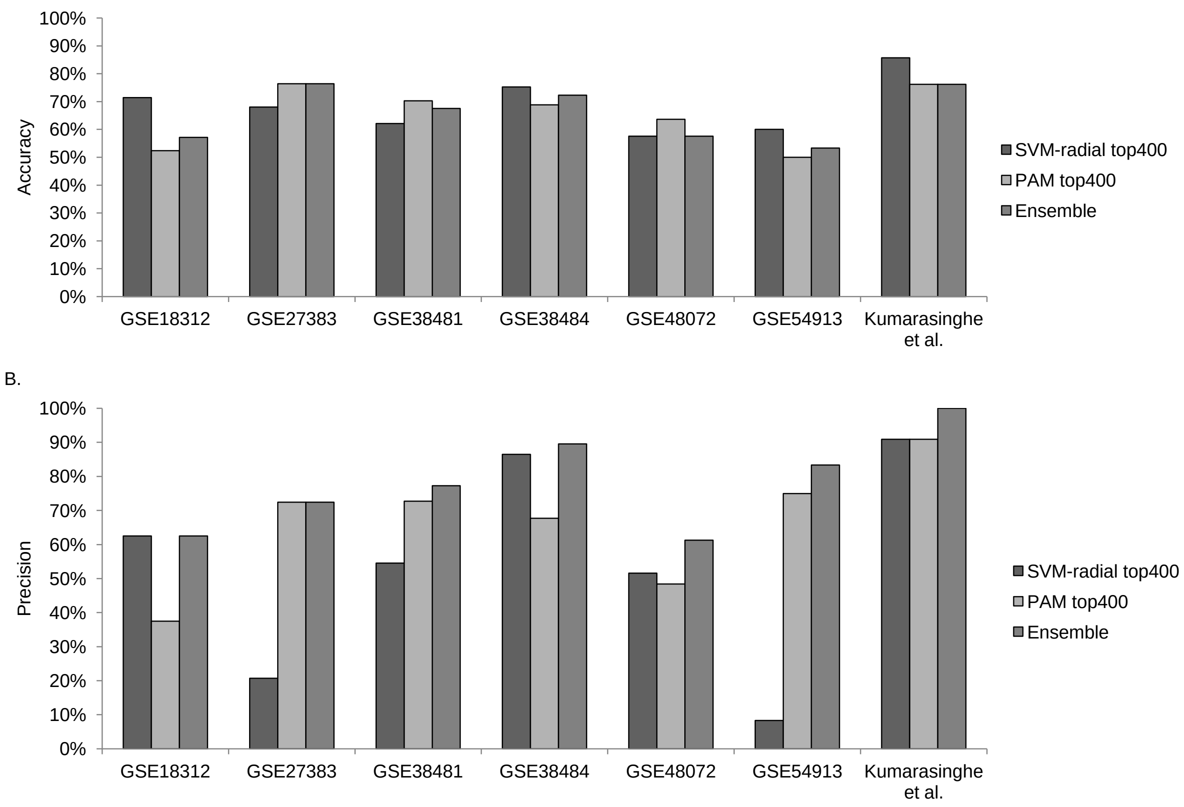


**Supplementary figure 10:** Test data prediction accuracy and precision of SVM-radial, PAM and ensemble model for each microarray dataset. The ensemble model with top 400 differentially expressed genes was evaluated for its performance across each dataset. A) No specific trend was observed for ML models in class prediction of the datasets irrespective of the tissue type, medication status and sample number. B) However, a consistent precision of ≥ 60% for all the dataset indicates a better performance ensemble model over SVM-radial and PAM.
