## Supplementary Table for "Ensemble learning for higher diagnostic precision in schizophrenia using peripheral blood gene expression profile"

**Supplementary Table 1:** Alignment and demographic details of all RNA-Seq samples

| **Sample ID** | **Alignment (%)** | **Diagnosis** |
| --- | --- | --- |
| 1001 | 84.75 | SCZ |
| 1002 | 95.31 | SCZ |
| 1003 | 85.85 | SCZ |
| 1005 | 95.53 | SCZ |
| 1006 | 83.61 | SCZ |
| 1007 | 80.16 | SCZ |
| 1008 | 94.10 | SCZ |
| 1009 | 79.92 | SCZ |
| 1010 | 88.43 | SCZ |
| 1011 | 80.18 | CNT |
| 1012 | 81.65 | CNT |
| 1013 | 85.97 | CNT |
| 1014 | 82.79 | CNT |
| 1016 | 84.63 | CNT |
| 1017 | 93.83 | CNT |
| 1018 | 95.11 | CNT |
| 1019 | 93.75 | CNT |
| 1020 | 81.67 | SCZ |
| 1021 | 94.80 | CNT |
| 1022 | 95.11 | CNT |

(Note: Sequences were aligned to the human genome (GENECODE hg38) using HISAT2.)
